# Effects of Different Mask Policies in 2020: A Comparative Analysis

**DOI:** 10.1101/2022.11.04.22281935

**Authors:** Xue-Jing Liu

**Author notes:** Correspondence concerning this article should be addressed to name, institution, postal address.

## Abstract

The research around the public’s usage of masks to prevent the spread of COVID-19 is developing quickly. In this work, we analyzed data from 50 nations to assess the long-term effectiveness of mask policies with different levels using the Poisson regression model and generalized linear mixed model. Over the long term, stricter obligatory mask regulations were linked to more stable patterns and slower increases in Covid-19 case occurrences. The mitigation of disease transmission by mask policies was shown to have substantial major impacts throughout the entire year of 2020, whereas the incidence of illness displays increasing trends over time under various policies. When compared to no mask policy deployment, mask policies might reduce incidence growth by 13.5% to 17.8%, although the incidence under every policy climbed 1.5% to 1.9% on average every ten days. The mask policy is effective in controlling illness, according to the bulk of the data shown above. This result confirms the mask policy’s importance as a governing approach in the context of the worldwide pandemic.

---

The sudden beginning and broad reach of the COVID-19 epidemic have led to significant adjustments in both individual behaviors and governmental practices. The government has advised or required mask use as one of the non-pharmaceutical preventative behaviors for disease control. On June 5, 2020, the World Health Organization (WHO, 2020) amended its recommendations and guidelines for the use of masks. Soon after, 175 nations released their own plans (Masks4All, 2020), which in total covered 99.78% of the world’s population. However, both globally and within a single nation, there have been variations in both government enforcement and civilian adherence to the laws over time. Its effectiveness in preventing disease transmission has also been questioned when combined with cultural diversity.

Planning and crisis management by the authorities will be made easier if we have a better understanding of the effectiveness of the mask policy in the context of a true pandemic scenario. Beyond the current crisis, our investigation also revealed details regarding the role mask policies play in the prevention of the spread of respiratory diseases and the extent to which they are effective over the long run. In the meanwhile, it offers a rare chance for public health education and prompts individuals to reconsider their own health and preventative measures.

## Literature Review

The covid-19 epidemic has sparked intense scholarly discussion and sped up the development of both theoretical and empirical literature. Prior researches have mostly examined the mask policies from the viewpoints of (1) the cultural, social, and psychological components of the regulations; (2) the compliance with the policies; and (3) the effectiveness in avoiding disease transmission.

The mask-covering has been a subject of ample discussions of cultural and social issues. Since the 1990s, Japanese have worn masks as a sign of civility during the winter (Burgess & Horii, 2012), and this is consistent with the “orientalist” description of Japanese culture as being characterized by its “groupism” and “collectivism” (Suzuki et al., 2010; Takano & Osaka, 1999). Numerous researches done in Asian nations have corroborated this idea, revealing that high rates of mask policy adherence were seen in Vietnam (99.5% at the beginning of March) (Nguyen et al., 2020), China (98% at the end of January) (Zhong et al., 2020), and Japan (70.1% at the end of March) (Muto et al., 2020). An international comparison (Zhao & Knobel, 2021) found that differences in subjective norm, risk-susceptibility, risk-severity, and conformity to authorities’ guidance explained why China had higher compliance with mask rules than Austria, Germany, and Switzerland. Based on social psychology, mask wearing was identified as a cultural activity that showed divergences in individualism-collectivism cultural frames, independent and interdependent self-construal, the tightness-looseness framework, American honor cultures and political orientation (Kemmelmeier & Jami, 2021).

Mask policy adherence is essential for containing the disease during the pandemic as a preventative measure (Fischer et al., 2021). The studies on the policy compliance suggested that behavioral recommendation by the government affected individuals’ protective behavior adoption by increased 12% mask wearing and 7% mask buying (Goldberg et al., 2020). While a research from Germany (Betsch et al., 2020) said that wearing a mask willingly would be seen as unfair and would heighten stigmatization, which would result in poor compliance. Contrarily, mandate in use by the authorities was thought to be efficient, fair, and socially responsible, and it can boost real compliance. After the required usage announcement in Australian city, the use of mask reached 97% and 100% respectively, based on photographs of public areas and interviews (Scott et al., 2021). The demographic features of the compliance rates were found to be greater among older adults, women, and residents of metropolitan regions (Guerra-Centeno et al., 2020; Haischer et al., 2020). A context analysis (He et al., 2021) further revealed that mask resistance was frequently motivated by physical discomfort, negative effects, lack of effectiveness, being unnecessary or inappropriate for some persons or in certain situations.

Studies on the benefits of mask policies typically focus on how they reduce the spread of disease. A Japanese research (Sugimura et al., 2021) reported a 0.4 relative risk of infection for mask users versus non-users. Additionally, a mandatory policy was related to a reduction of 47% daily growth rate in reported cases (Mitze et al., 2020), and mandate use among employees was related to a 10% reduction in weekly growth rate of cases and deaths (Chernozhukov et al., 2021). Some studies further pinpointed the superiority of timing in the implementation of policy. Comparing with other governmental strategies, early adoption of mandate mask wearing reported the strongest short and long-term effects on curbing infections (An et al., 2021) Specifically, early policy adopters reported a magnitude of 1% in cases reduction, while the later adopters reported a 0.44% decrease comparing non-adopter in a statewide comparison study (Rebeiro et al., 2021).

Among these studies, most of them had illustrated the short- or medium-term effectiveness of the mask policy by documenting a rapid shift in their implementation. While the unanticipated extension of the pandemic’s length forces governments to take a long-term strategy and analyze the supporting data. Moreover, most of the studies had a local focus or had data collected for only one nation. It would be ideal to evaluate policies thoroughly. Even though research policy that focuses on a single location could offer more thorough material for reference, a worldwide landscape is still required. Antecedent studies came to the conclusion that the policy’s efficacy was mostly estimated from before-after or voluntary-mandatory comparisons. They disregarded the policy descriptions’ more specific criteria. We want to give further information regarding the differences in their efficiency for segmentable policy needs as a result of the data that is already accessible.

In this study, (1) We investigated the mask policy for the entire 2020 year in order to assess its effectiveness over the long term; (2) We investigated the mask policy by including 50 countries that reported the highest cumulative cases globally in order to provide a conclusion from a comprehensive standpoint; and (3) We distinguished the corresponding effects of mask policies with 5 levels of different stringencies, ranging from policy 0 (no policy) to policy 4. (mask mandates for all places).

## Methodology

### Date Sources

We adopted daily mask policies and daily Stringency Index data for each country from 1^st^ January to 31^st^ December, 2020 from the Oxford COVID-19 Government Response Tracker (Hale et al., 2020). The policies were divided into five categories (0-4) that represented various demands for national authorities to wear masks: 0 = No policy; 1=Masks are recommended; 2= Mask are required in some specified shared/public spaces outside the home with other people present, or some situations when social distancing not possible; 3= Masks are required in all shared/public spaces outside the home with other people present or all situations when social distancing not possible; 4=Masks are required outside the home at all times regardless of location or presence of other people. Policies with higher number indicate more stringent gradients. In accordance with their specifications, we further classified the policies as follows: policy 0 is no policy or free policy; policy 1 is a voluntary mask policy; policy 2, policy 3, and policy 4 are required mask policies, and their requirements are more stringent as the number increases.

The country’s Stringency Index is composited by measuring nice metrics of governmental policies other than mask policy during the pandemic period: closure of schools, workplaces, public events, bans on public gatherings, suspension of public transportation, mandates for people to stay at home, public awareness campaigns, restrictions on internal movement, and international travel controls.

50 nations that reported the highest number of cumulative cases at the end of 2020 were included in this the study. The research periods for each nation began when it reported the first confirmative case until the end of the 2020. Considering disease incubation periods and lay-time for an intervention to take effect, we believed that the policy less than 15 days was unable to go into effect among the population. Therefore, we recorded a mask policy as the previous one level if it had been in place for less than 15 days.

We obtained the daily new cases of covid-19 between 1st January 2020 and 31st December 2020 at the country level from the World Health Organization (WHO, 2020). We estimated the daily 7-day smoothed confirmed new cases using the 7 days moving average approach to remove the effect of the date reporting due to administrative reasons (weekend, lab shut, etc.). The statistic began with the day when its 7-day smoothed cases no smaller than 1 until December 31, 2020.

### Statistical Analysis

The primary outcome of this study was the daily changes in incidence rate for every 10 days. The 7-day smoothed new confirmed cases were used to fit the Poisson regression model, controlled by the daily countries’ stringency index to reduce confounding. At the same time, we conducted the sensitivity analysis to check the robustness of this primary analysis by estimating the daily changes of incidence calculating every 15 days and every 30 days data respectively.

The secondary outcome were the effects of the policy on the incidence rate. The data were analyzed by fitting a generalized linear mixed model (GLMM) (Barr et al., 2013) using the normal distribution with identity link function. First, the fixed-effect model was built, and the period following the adoption of the policy was examined as a continuous independent variable, and the dependent variable was the incidence rate determined using Poisson regression.

We also investigated the interaction impact of policy and time to determine its predictive power for longitudinal changes in the dependent incidence. The random effect model was then tested by adding an intercept and enabled the individual policy periods starting with random incidence. We also incorporated time as a predictor of slope change and allowed specific policy periods to vary in a longitudinal trajectory. The results of the GLMM are reported as estimated coefficient with 95% confidence intervals. A two-tailed p value less than 0.05 was considered statistically significant.

## Results

### Policies Characteristics

Countries reported the national first new case around March 2020, and additional confirmed cases continued to mount up after that. In total, 100 mask policy periods have been applied throughout the 50 nations in 2020. As shown in table 1, 32 (64%) counties implemented policy 3 with 38 periods, and almost half of the nations had carried out policy 2 and 4, respectively.

**Table 1.**
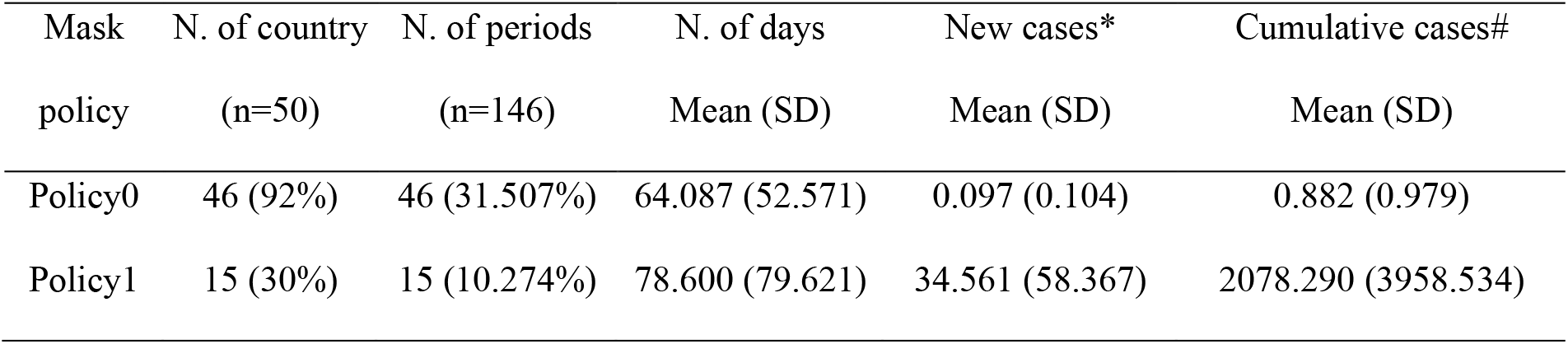
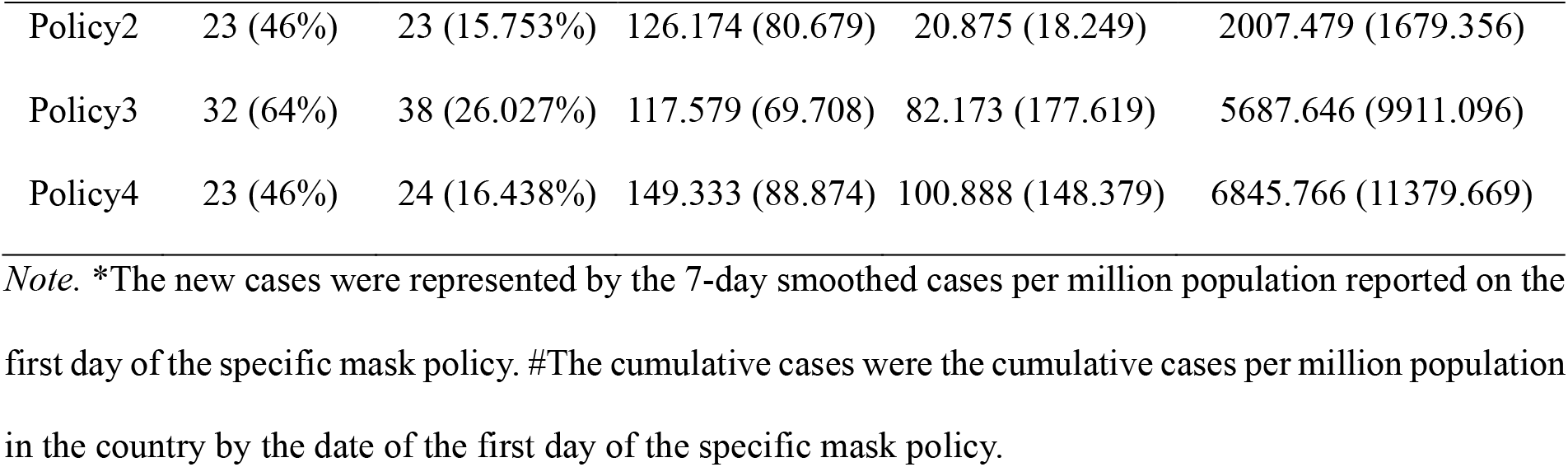
Characteristics of the Mask Policy from 50 Countries.

| Mask policy | N. of country<br>(n=50) | N. of periods<br>(n=146) | N. of days<br>Mean (SD) | New cases*<br>Mean (SD) | Cumulative cases#<br>Mean (SD) |
| --- | --- | --- | --- | --- | --- |
| Policy0 | 46 (92%) | 46 (31.507%) | 64.087 (52.571) | 0.097 (0.104) | 0.882 (0.979) |
| Policy1 | 15 (30%) | 15 (10.274%) | 78.600 (79.621) | 34.561 (58.367) | 2078.290 (3958.534) |
| Policy2 | 23 (46%) | 23 (15.753%) | 126.174 (80.679) | 20.875 (18.249) | 2007.479 (1679.356) |
| Policy3 | 32 (64%) | 38 (26.027%) | 117.579 (69.708) | 82.173 (177.619) | 5687.646 (9911.096) |
| Policy4 | 23 (46%) | 24 (16.438%) | 149.333 (88.874) | 100.888 (148.379) | 6845.766 (11379.669) |

*Note.* \*The new cases were represented by the 7-day smoothed cases per million population reported on the first day of the specific mask policy. #The cumulative cases were the cumulative cases per million population in the country by the date of the first day of the specific mask policy.

The average length of policy 4 was 149.333 days, followed by policy 2 (126.174 days on average), and the average length of policy-free (policy 0) was 64.087 days. These countries reported an average of 101 new cases per million population and 6846 cumulative cases per million of the population when they implemented the policy 4. They reported an average of 82 new cases and 5688 cumulative cases per million population when implemented policy 3; 34.561 new cases and 2078.29 cumulative cases when implementing the policy 1.

China first advised individuals to wear masks on January 12th, then made it mandatory while entering all shared/public venues (policy 3) on January 22nd. Following that, France advised mask use on February 28th, Japan on March 1st, Brazil on March 7th, and the United States on March 10th. On March 18th, Czechia became the first country outside of China to establish an obligatory policy (Policy 3). Four days later, Brazil issued an obligatory regulation forcing individuals to wear masks in some shared/public locations (policy 2). On April 4th, Italy and the United Arab Emirates introduced Policy 4, which required masks to be worn at all times when individuals were outside of their homes. Indonesia (April 6th), Colombia (April 7th), Morocco (April 7th), India (April 9th), and Brazil (April 11th) quickly issued the regulation. Appendix Table A has more extensive information on 50 nations.

Specifically, Figure 1 shows the situation of mask policies, daily cases, and regulation stringency for each country that included in this study.

**Figure 1.**
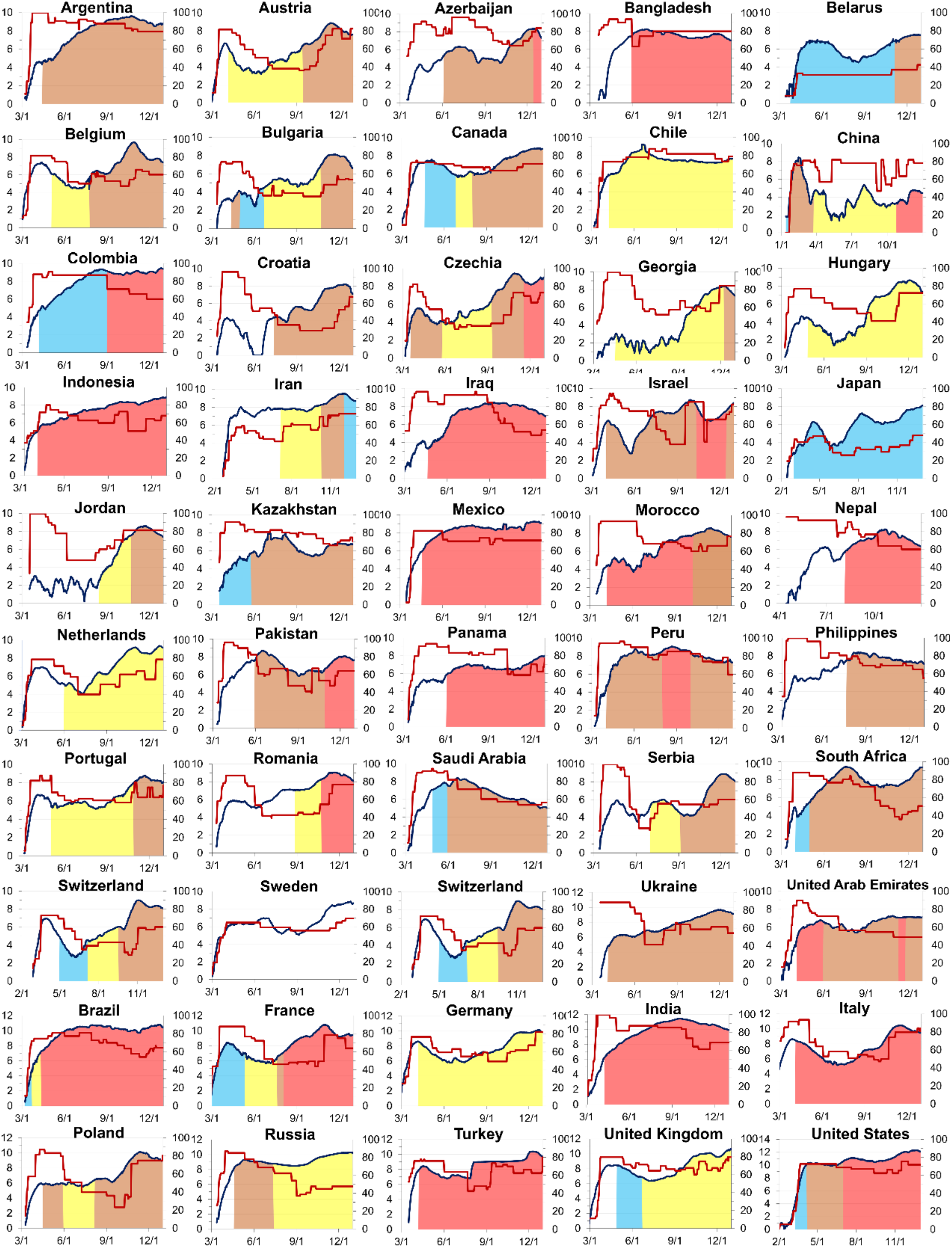
Mask Policy, Covid-19 Cases, and Stringency Index for 50 Countries. *Note*. The daily covid-19 cases are represented by the natural logarithm of daily 7-days smoothed newly confirmed cases, which showed as the black curves with the scales on the left side (ranging from 0-12, and 0-14 for the United States) for reference. The colorings under the dark curves in each graph show the different mask policies during relevant periods. The transparent areas indicate policy 0, and blue for level 1, yellow for level 2, orange for level 3, and red for level 4. The stringency index is represented by red curves along the study periods with the scales on the right side (range from 0-100).

Several countries followed a single policy throughout the year. For example, Sweden did not implement any mask policies, and Japan only implemented policy 1. Four countries (Chile, Hungary, the Netherlands, and Germany) only implemented policy 2, three countries (Argentina, the Philippines, and Ukraine) only implemented policy 3, and nine countries (Bangladesh, Indonesia, Iraq, Mexico, Nepal, Panama, India, Italy, and Turkey) implemented policy 4 for the majority of 2020. Other nations employed combinate policies in 2020, and the majority of them (29 out of 30) had ever raised the stringency of the rules, with 22 showing a rising tendency in stringency without revising down.

### Result for Poisson Analysis

The Figure 2 shows the Poisson regression result for daily incidence and changes under different mask policies

**Figure 2.**
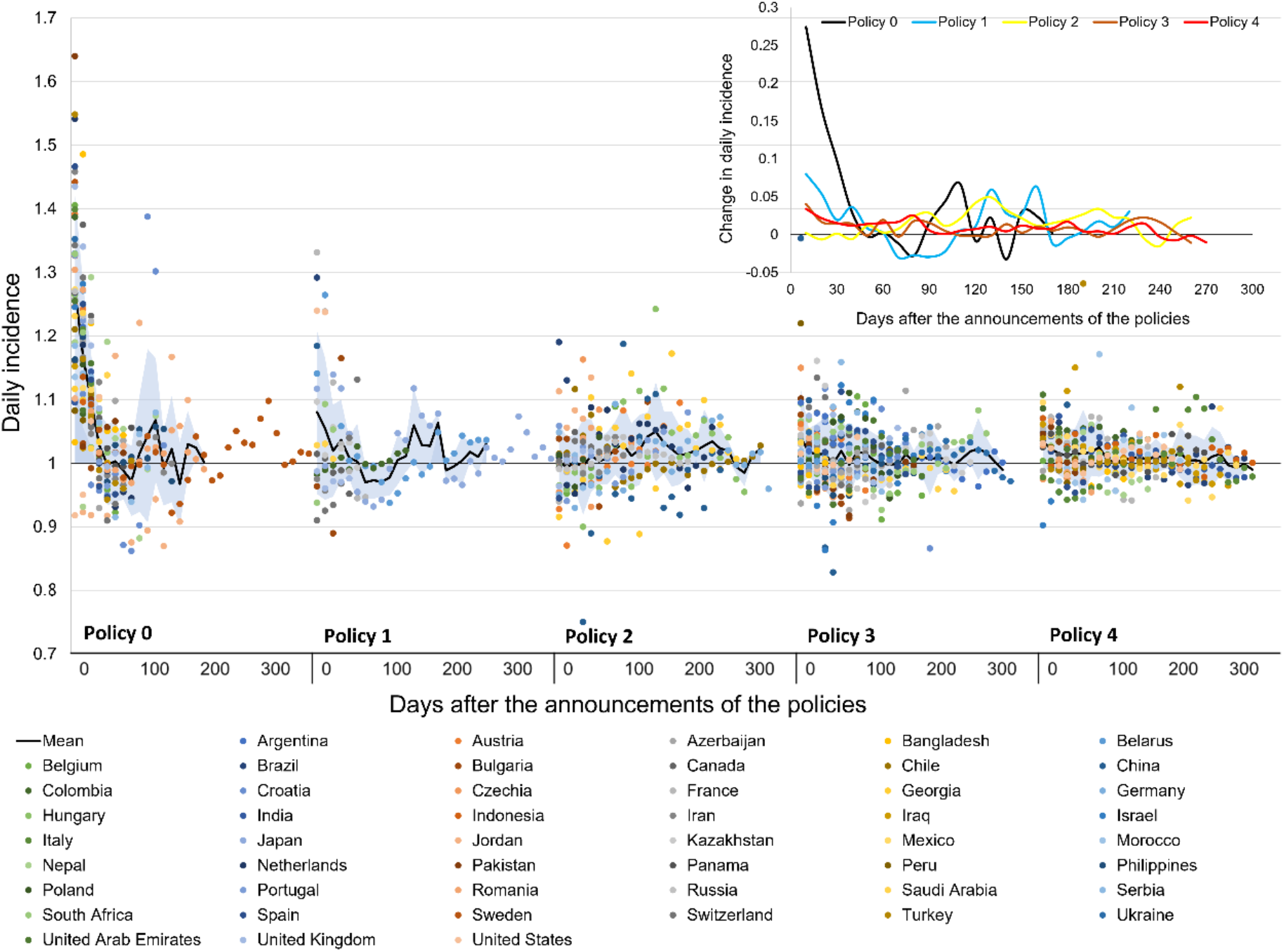
The Daily Incidence along the Time under Different Mask Policies *Note*. Dots represent countries’ daily incidence, black lines represent the means of the daily incidence at a specific time, and shaded areas represent mean±SD. The curves in the upper-right side depicted trends of incidence changes under different policies for the study period.

As shown in Figure 2, policies 2, 3, and 4 were adopted by a greater number of nations and were likely to be maintained for a longer period of time. incidence changes rate for most policies decreased from day 10 to day 50, although they remained greater than 1. Changes of incidences under policies 0, 1, and 2 appeared to fluctuate a lot from day 60 to day 170, ranging from -3.270% to 6.713%. Policy 3 and policy 4 demonstrated a consistent rise in incidence with a lower magnitude throughout the year.

Without any mask strategy, the incidence of new cases increased by 4.076% per day on average in one year, as seen in Table 2. Changes in policy 1 and policy 2 are comparable, with roughly a 1.5% rise every day. At the same time, policies 3 and 4 had similar daily incidence increase rates of 0.9% each day on average.

**Table 2.**
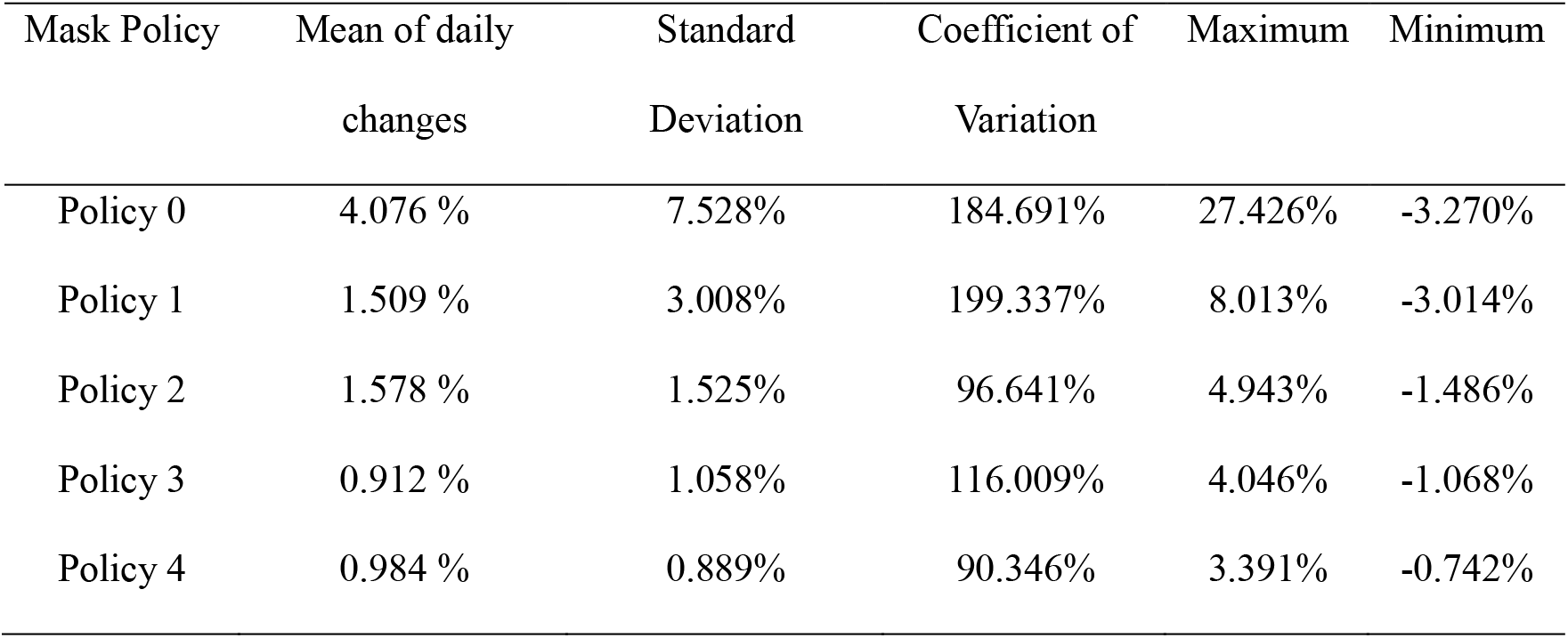
Statistics of Average Incidence Changes under Different Mask Policies.

| Mask Policy | Mean of daily changes | Standard Deviation | Coefficient of Variation | Maximum | Minimum |
| --- | --- | --- | --- | --- | --- |
| Policy 0 | 4.076 % | 7.528% | 184.691% | 27.426% | -3.270% |
| Policy 1 | 1.509 % | 3.008% | 199.337% | 8.013% | -3.014% |
| Policy 2 | 1.578 % | 1.525% | 96.641% | 4.943% | -1.486% |
| Policy 3 | 0.912 % | 1.058% | 116.009% | 4.046% | -1.068% |
| Policy 4 | 0.984 % | 0.889% | 90.346% | 3.391% | -0.742% |

### Result for GLMM analysis

The GLMM with normal distribution revealed significant main effects of mask policy, χ^2^(4) = 102.558, p < 0.000, and a linear mixed model showed a reliable main effect of time, χ^2^(1) = 68.736, p < 0.000, as well as the interaction of policy and time, χ^2^(4) =39.769, p < 0.000. The results of GLMM analysis are showed in the table 2.

These results revealed that the time and conducting policies with enhanced stringencies were independently associated with reductions in incidence for the whole year of 2020. As a main effect in the incidence drop, time was related to 1.7% decrease every 10 days. All policies demonstrated decreased incidence in 2020 when using policy 0 as the reference group. The incidence under policy 2 showed a statistically significant reduction of 17.8%, followed by policies 3 and 4 with reductions of 15.6% and policy 1 with reductions of 13.5%.

When taking into account the interplay between time and policy levels, we used the interaction term of policy 0 and time as the reference group and displayed the incidence changes over time under each unique policy level. Results revealed that incidents have also grown by 1.15 to 1.16 percent every 10 days under policies 1, 3, and 4, and by 1.9% under policy 2.

The outcomes of the random effect model supported time as a random effect factor but did not support a random intercept. It suggested the existence of statistically significant heterogeneity in the trends of changes under different policies, but the estimation was small.

**Table 2.** The Main Effects of the Fixed Model and the Random Model.

|  | Coefficient | P value | 95% CI |  |
| --- | --- | --- | --- | --- |
|  |  |  | Lower | Upper |
| The main effects of the fixed model |  |  |  |  |
| Intercept | 1.178 | <0.001 | 1.165 | 1.192 |
| Time | -0.017 | <0.001 | -0.020 | -0.015 |
| Main effects of the mask policy |  |  |  |  |
| Policy 0 | Reference |  |  |  |
| Policy 1 | -0.135 | <0.001 | -0.160 | -0.110 |
| Policy 2 | -0.178 | <0.001 | -0.198 | -0.158 |
| Policy 3 | -0.156 | <0.001 | -0.175 | -0.138 |
| Policy 4 | -0.156 | <0.001 | -0.175 | -0.136 |
| Main effects of the interaction of time and policy |  |  |  |  |
| Time*Policy 0 | Reference |  |  |  |
| Time*Policy 1 | 0.015 | <0.001 | 0.010 | 0.019 |
| Time*Policy 2 | 0.019 | <0.001 | 0.016 | 0.022 |
| Time*Policy 3 | 0.016 | <0.001 | 0.013 | 0.019 |
| Time*Policy 4 | 0.016 | <0.001 | 0.013 | 0.019 |
| The effects of the random model |  |  |  |  |
| Intercept | 0.000 | 0.593 | 0.000 | 0.003 |
| Time | 0.000 | 0.032 | 0.000 | 0.000 |

*Note.* The probability of the distribution is normal, the dependent variable of the analysis is the daily incidence rate.

## Discussion

In this study, we used data from 2020 from 50 nations to give conclusions from a global perspective on the efficacy of various mask regulations. The findings suggested that mask policies contributed to a long-term public health plan to address the pandemic problem in the dimensions of both national and international.

The dates that different countries enacted policies and their preferred policy levels varied. Most nations had maintained a stricter policy or increased it in response to the rising incidences. When the number of new cases and accumulated cases was large, governments typically implemented regulations with higher stringencies, while these cases on the announcement of policy 1 were slightly higher than that for policy 2. A possible explanation for this is that both policies are considered as the first policy at the start of a national epidemic. The decisions they make in this case are heavily depend on the judgments of each particular government, thus this preference could introduce moderation in the differences between them.

The daily incidence rate was often higher than 0.9 throughout 2020, and all levels of mask rules were substantially linked with decreases in annual incidence changes when compared to mask-free policy. Compared to policy 1 and policy 2, policy 3 and policy 4 exhibited lower and more steady daily growth rates. Additionally, the sensitivity analysis produced similar results, with the exception that the policy 1 reported a lower incidence growth while a large coefficient of deviation than that for policy 3 using 30 days data. This finding corroborated the results achieved by several studies that respectively looked for policies’ impact on the reduction in transmission growth (Rader et al., 2021), efficiencies of voluntary mask policy and mandatory policy (Lyu & Wehby, 2020) and it further supported the idea of mask policy use in transmission control.

The GLMM model analysis demonstrated that each strategy had an approximately 13.5% to 17.8% reduction in the accelerated pace over the course of the whole year when compared to policy-free. When considering the longitudinal trends, it was found that for each strategy, the incidence rate fluctuated between 1.5% and 1.9% every ten days. This evidence again supported a constructive association between mask policies and transmission control, but it also highlighted the point that these effects had gradually weakened over time. Potential explanation for these may be mask wearing decreasing personal vigilance for protection and leading to less other precautionary behaviors. These behaviors include frequent face touching due to the physical discomfort that masks cause, spending more time in shared spaces with others present, and going into a crowed place. The authorities may also overestimate the efficiency of mask wearing, thus approved for reopen the public facilities (restaurant, school, university, etc.) and resumed public activities.

Results from Poisson analysis and GGLM analysis in current study all indicated that the policy 4, which added more requirements based on the policy 3, did not imply a boost to the efficacy. The hazy areas in policy definition and performance may in part contribute to this outcome’s explanation. The differences between policy 3 and policy 4 can occasionally be small and insignificant when combined with other non-pharmaceutical strategies, such as shelter in place, closure of schools and public facilities, work from home, cancellation of gatherings, etc. Particularly when people are unclear about how the policy should be applied (e.g., “public spaces with or without other people present” and “when social distancing do or do not possible”), they may interpret the rule based on their own judgement of the likelihood of infection and act accordingly. This might skew the estimations of the efficiency for the mandatory policies (policy 2, 3 and 4) we calculated here.

There are some limitations in this study. Firstly, it failed to control all the confounders. We had included the social stringency index as a covariate to constrain other interventions measures’ bias on the changes of incidence. This does not, however, rule out other variables from a personal, environmental, national, or international perspectives from playing a role in the variations in incidence.

Secondly, the analysis only considered nations that had the highest cumulative case counts for 2020. It did not consider data from nations with smaller populations but were highly infectious as well as nations with moderate and low cumulated cases, thus the study is not sufficiently representative. In order to fully comprehend the mask policy, more research on these nations is required.

Thirdly, we assumed that the policy compliance rate was the same for every country as well as during the whole study period, or different rates contribute to equivalent effectiveness. Obviously, reality can challenge these assumptions easily. A model (Stutt et al., 2020) suggested that even with a 30% or 50% deficiency in mask effectiveness, 100% policy adherence would produce the best results. Additionally, two studies (Yan et al., 2019) noted that disease might be gradually wiped out with at least 80% adherence to universal usage. In actuality, it was discovered that the rates varied greatly in terms of culture, nation, ethnicity, season, and other demographic parameters. A survey (YouGov, 2020). reveals the variation in mask behavior between nations. Some Asian countries reached an 80% compliance rate around the middle of April, three European countries (Italy, Span, France) did that on June 20th, and the United States and Canada reached this level at the end of August, while other counties failed to do that. It could be anticipated that considering them will depreciate the effects of policies we had found in this study. Therefore, a multivariable analysis including these factors is desirable in the future.

Fourthly, the results in this study supported the notion that the nation and globe may be beneficial by the mask use, while we failed to consider the types of masks use and their difference in virus preventing. Early policies were based on the temporal research conclusions that the main transmission through droplet and propulsion of large particles could be prevented by face shields, thus cloth mask and surgical masks were normally suggested by the policies. However, this knowledge was soon challenged by updated findings that smaller airborne particles are infectious and cannot be prevented by a cloth or surgical mask, and a respirator is more desirable (Lindsley et al., 2021).

Take these renewals into consideration, results in this article may underestimate its true effects. By this way, we could believe that mask policy could be a more effective measure to prevent infections if sufficient masks are provided with increased efficiency. In conclusion, the results are providing evidence that mask policy, especially mandatory mask policy, could be considered as an essential tool for transmission containment in the strategy pocket in the covid-19 pandemic. While more evidence is still needed for people to better understand the face-covering behavior and mask policy.

## Data Availability

All data produced are available online at the reference

