## Appendix Table A for "Effects of Different Mask Policies in 2020: A Comparative Analysis"

*Characteristics of Incidence and Mask Policies in Different Countries*

| | Policy 1 | | | Policy 2 | | | Policy 3 | | | Policy 4 | | |
|---|---|---|---|---|---|---|---|---|---|---|---|---|
| Country | Starting Date | New Cases* | cumulative Cases# | Starting Date | New Cases* | cumulative Cases# | Starting Date | New Cases* | cumulative Cases# | Starting Date | New Cases* | cumulative Cases# |
| Argentina | | | | | | | 4/14 | 2.206 | 49.828 | | | |
| Austria | | | | 4/6 | 48.505 | 1368.139 | 9/14 | 63.384 | 3732.569 | | | |
| Azerbaijan | | | | | | | 6/3 | 19.599 | 558.428 | 12/14 | 410.008 | 16906.994 |
| Bangladesh | | | | | | | | | | 5/30 | 10.869 | 270.861 |
| Belarus | 3/25 | 0.680 | 8.572 | | | | 11/5 | 102.320 | 10827.548 | | | |
| Belgium | | | | 5/4 | 38.150 | 4424.907 | 7/25 | 27.981 | 5751.179 | | | |
| Brazil | 3/7 | 0.008 | 0.080 | 3/22 | 0.524 | 4.253 | | | | 4/11 | 6.685 | 84.009 |
| Bulgaria | 5/1 | 8.409 | 216.739 | 6/22 | 12.644 | 561.996 | 4/12 | 3.248 | 95.129 | | | |
| | | | | | | | 10/23 | 152.614 | 5027.024 | | | |
| Canada | 4/20 | 38.634 | 898.438 | 6/26 | 9.043 | 2708.960 | 8/1 | 11.836 | 3068.161 | | | |
| Chile | | | | 4/8 | 17.771 | 267.626 | | | | | | |
| China | 1/12 | 0.004 | 0.031 | 3/24 | 0.063 | 56.795 | 1/22 | 0.039 | 0.307 | 10/24 | 0.019 | 63.677 |
| Colombia | 9/1 | 187.520 | 11947.788 | | | | | | | 4/7 | 2.198 | 29.185 |
| Croatia | | | | | | | 7/13 | 19.870 | 906.640 | | | |
| Czechia | | | | 5/25 | 6.470 | 838.829 | 3/18 | 5.163 | 42.114 | 11/18 | 532.290 | 43874.016 |
| | | | | | | | 9/10 | 70.248 | 2899.716 | | | |
| France | 2/28 | 0.057 | 0.582 | 5/11 | 16.119 | 2099.979 | 7/20 | 10.385 | 2582.885 | 8/3 | 19.805 | 2820.685 |
| Georgia | | | | 4/17 | 5.014 | 92.751 | 12/8 | 1085.296 | 42527.425 | | | |
| Germany | | | | 4/6 | 64.951 | 1138.536 | | | | | | |
| Hungary | | | | 4/27 | 8.858 | 267.382 | | | | | | |
| India | | | | | | | | | | 4/9 | 0.404 | 4.250 |

| | | | | | | | | | | | | |
|---|---|---|---|---|---|---|---|---|---|---|---|---|
| Indonesia | | | | | | | | | | 4/6 | 0.563 | 9.107 |
| Iran | 12/3 | 161.896 | 11781.608 | 7/5 | 30.101 | 2832.119 | 10/10 | 47.252 | 5862.135 | | | |
| Iraq | | | | | | | | | | 4/20 | 0.664 | 38.262 |
| Israel | | | | | | | 4/1 | 60.721 | 662.928 | 10/12 | 394.347 | 33724.663 |
| | | | | | | | 12/15 | 211.541 | 41624.810 | | | |
| Italy | | | | | | | | | | 4/4 | 78.749 | 1981.862 |
| Japan | 3/1 | 0.121 | 1.890 | | | | | | | | | |
| Jordan | | | | 8/15 | 1.288 | 136.919 | 10/23 | 187.113 | 4551.641 | | | |
| Kazakhstan | 3/17 | 0.251 | 1.757 | | | | 5/25 | 16.875 | 477.666 | | | |
| Mexico | | | | | | | | | | 4/17 | 2.954 | 45.349 |
| Morocco | | | | | | | 10/10 | 69.020 | 3966.290 | 4/7 | 2.194 | 30.913 |
| Nepal | | | | | | | | | | 8/6 | 10.801 | 746.478 |
| Netherlands | | | | 6/1 | 10.080 | 2696.022 | | | | | | |
| Pakistan | | | | | | | 5/30 | 9.067 | 300.857 | 10/29 | 3.533 | 1494.846 |
| Panama | | | | | | | | | | 6/2 | 83.997 | 3120.215 |
| Peru | | | | | | | 4/2 | 3.652 | 40.125 | 8/1 | 157.693 | 12358.783 |
| | | | | | | | 10/1 | 152.606 | 24620.029 | | | |
| Philippines | | | | | | | 7/22 | 17.347 | 645.768 | | | |
| Poland | | | | 5/30 | 9.572 | 611.812 | 4/16 | 8.972 | 200.335 | | | |
| | | | | | | | 8/6 | 16.506 | 1289.125 | | | |
| Portugal | | | | 5/3 | 21.113 | 2470.405 | 10/28 | 289.953 | 12203.153 | | | |
| Romania | | | | 8/27 | 59.623 | 4244.064 | | | | 10/23 | 204.316 | 10188.541 |
| Russia | | | | 7/13 | 44.870 | 5027.592 | 4/19 | 26.512 | 293.646 | | | |
| Saudi Arabia | 4/28 | 34.169 | 540.331 | | | | 5/30 | 57.641 | 2348.662 | | | |
| Serbia | | | | 7/1 | 24.067 | 1666.863 | 9/4 | 9.434 | 3625.347 | | | |
| South Africa | 4/1 | 2.226 | 26.390 | | | | 5/1 | 4.720 | 110.144 | | | |

| | | | | | | | | | | | | |
|---|---|---|---|---|---|---|---|---|---|---|---|---|
| Spain | | | | 5/4 | 25.944 | 4838.842 | 5/21 | 13.334 | 5090.239 | 10/13 | 237.189 | 19736.785 |
| Sweden | | | | | | | | | | | | |
| Switzerland | 4/30 | 15.995 | 3429.497 | 7/6 | 11.290 | 3731.648 | 9/17 | 49.883 | 5666.683 | | | |
| Turkey | | | | | | | | | | 6/7 | 10.358 | 2006.401 |
| Ukraine | | | | | | | 4/4 | 2.868 | 25.061 | | | |
| United Arab | | | | | | | 5/31 | 74.993 | 3427.161 | 4/4 | 12.407 | 127.801 |
| Emirates | | | | | | | 11/25 | 123.583 | 16315.313 | 11/10 | 117.690 | 14487.682 |
| United Kingdom | 4/28 | 68.196 | 2318.386 | 6/22 | 14.072 | 4085.569 | 8/17 | 16.071 | 4709.041 | | | |
| United States | 3/10 | 0.246 | 2.260 | | | | 4/6 | 78.734 | 5.435 | 7/1 | 121.573 | 147.029 |
| N | | 15 | | | 23 | | | 38 | | | 24 | |
| Mean | | 34.561 | 2078.290 | | 20.875 | 2007.479 | | 82.173 | 5687.646 | | 100.888 | 6845.766 |
| (SD) | | (58.367) | (3958.534) | | (18.249) | (1679.356) | | (177.619) | (9911.096) | | (148.379) | (11379.669) |

Note. *The new cases were represented by the 7-day smoothed cases per million population reported on the first day of the specific mask policy. #The cumulative cases were the cumulative cases per million population in the country by the date of the first day of the specific mask policy.
